# Rapid onset of chronic urticaria after Moderna COVID-19 booster vaccine

**DOI:** 10.1101/2022.11.03.22281730

**Authors:** Olivier Duperrex, Francesco Tommasini, Yannick D. Muller

## Abstract

A substantial number of patients with newly onset of chronic urticaria was observed rapidly after the booster vaccine against COVID-19. This observation was unprecedented compared to the primary series of vaccination. To address this concern, we initiated an online observational study with the help of local allergists. We found a striking association between the booster dose, the Moderna vaccine, and the new onset of chronic urticaria within the following 10 days. These data were confirmed when reviewing all cases of CSU related to COVID-19 vaccination reported to Swissmedic, the Swiss regulatory agency. These data should not discourage patients from being vaccinated, as this vaccination campaign has been instrumental in reducing COVID-19 burden and preventing millions of deaths. Yet, there is an urgent need to establish appropriate guidelines and monitor this adverse event more closely, considering that a fourth dose is currently being administered.

## Introduction

The mRNA-based vaccines Spikevax from Moderna (hereafter referred to as Moderna) and Comirnaty from Pfizer-BioNTech (hereafter Pfizer) are the most widely distributed vaccines in Switzerland[1]. The immunization protocol consists of a primary series of two injections 3 to 4 weeks apart and a booster dose 4-6 months later. The booster was approved end of November 2021 by the Swiss regulatory agency (Swissmedic) for the ≥18-year-old population. Since then, we have observed a substantial number of chronic spontaneous urticaria (CSU) rapidly after the booster dose [2, 3]. CSU is defined as recurrent wheals, angioedema, or both for a duration of >6 weeks, frequently associated with a substantial impact on the health-related quality of life [4].

## Methods

As a collaborative effort with local allergists, we started an online observational study to better characterize individuals with newly diagnosed CSU possibly related to COVID-19 vaccines in the Canton of Vaud (CSU-VD). Among the 97 first patients, 80 accepted to participate in an online survey. In parallel, we reviewed all cases of CSU related to COVID-19 vaccination reported to Swissmedic (CSU-CH, n=782) (data kindly shared respecting data protection by I. Scholz, Pharmacovigilance Unit). We compared these data to the general population immunized with a booster in the Canton de Vaud (hereafter referred to as VD-booster, n=298,813) and in Switzerland (hereafter referred to as Booster CH, n=3,278,808). The “Commission cantonale (Prof. Pierre-André Michaud) d’éthique de la recherche sur l’être humain” CER-VD (BASEC 2021-00735, Lausanne, Switzerland, https://www.cer-vd.ch/) approved the study.

## Results

In 90% (CSU-VD) and 81% (CSU-CH) of the cases, CSU started after the booster. The median time between the vaccination and CSU onset was ten days. Strikingly, 9 out of 10 CSU cases were associated to Moderna vaccine (Table 1). At data collection, 31% patients reported a diagnosis of COVID infection. In those individuals, the median time between the vaccination and CSU onset was 51 days. The overall incidence rate of CSU after a COVID-19 booster per 100,000 persons immunized with a booster was similar in Vaud (24) and Switzerland (19, Table 2). Compared to Pfizer, the relative risk of developing CSU after Moderna was 21 (95% CI 6.5 - 66) and 16 (11 – 24) respectively (Table 2).

**Table 1.**
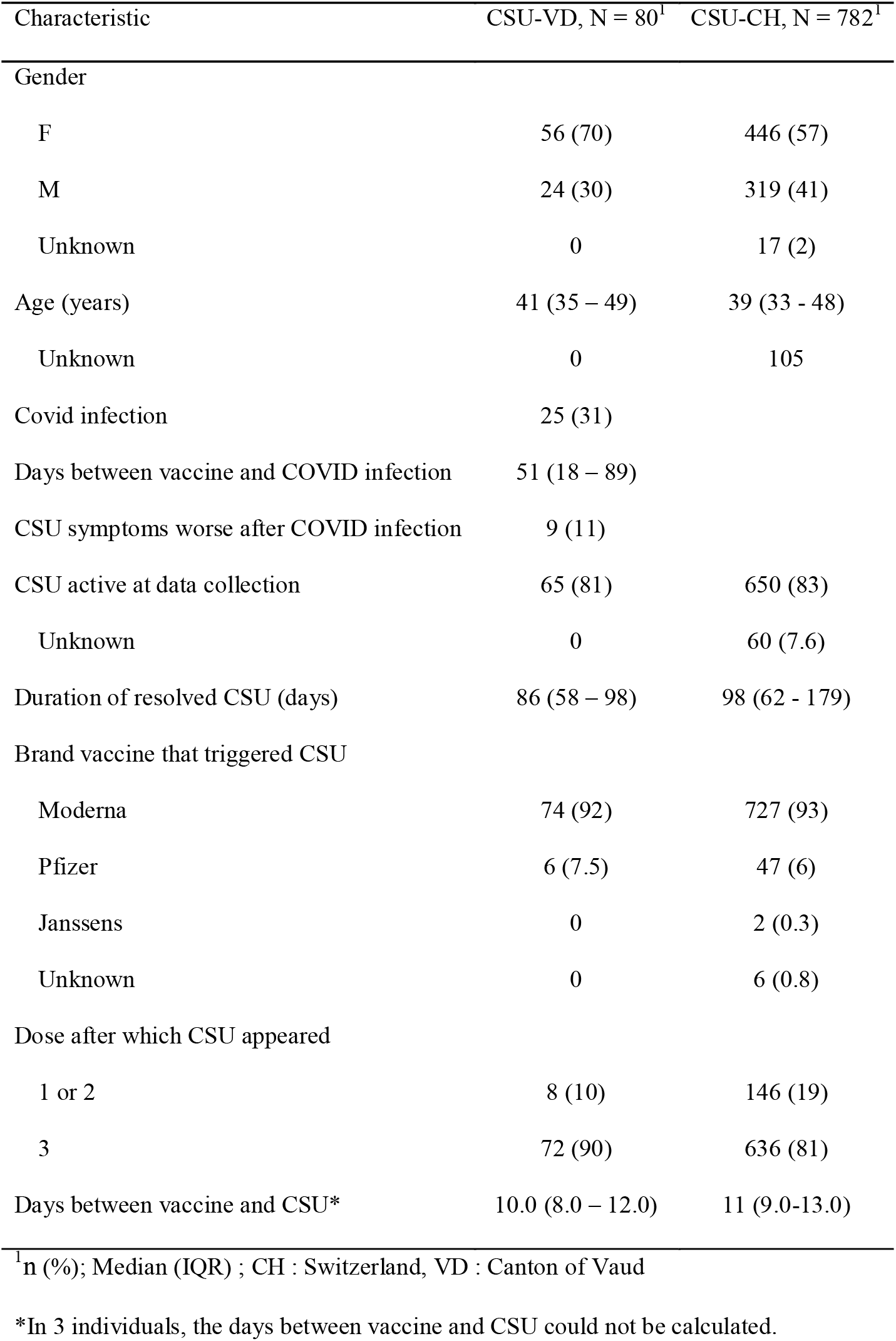
Chronic urticaria (CSU) after COVID-19 Vaccine - All CSU cases for Switzerland (CH) and Vaud (VD) [from 2021-01-21 to 2022-08-31]

| Characteristic | CSU-VD, N = 80<sup>1</sup> | CSU-CH, N = 782<sup>1</sup> |
| --- | --- | --- |
| Gender |  |  |
| F | 56 (70) | 446 (57) |
| M | 24 (30) | 319 (41) |
| Unknown | 0 | 17 (2) |
| Age (years) | 41 (35 – 49) | 39 (33 - 48) |
| Unknown | 0 | 105 |
| Covid infection | 25 (31) |  |
| Days between vaccine and COVID infection | 51 (18 – 89) |  |
| CSU symptoms worse after COVID infection | 9 (11) |  |
| CSU active at data collection | 65 (81) | 650 (83) |
| Unknown | 0 | 60 (7.6) |
| Duration of resolved CSU (days) | 86 (58 – 98) | 98 (62 - 179) |
| Brand vaccine that triggered CSU |  |  |
| Moderna | 74 (92) | 727 (93) |
| Pfizer | 6 (7.5) | 47 (6) |
| Janssens | 0 | 2 (0.3) |
| Unknown | 0 | 6 (0.8) |
| Dose after which CSU appeared |  |  |
| 1 or 2 | 8 (10) | 146 (19) |
| 3 | 72 (90) | 636 (81) |
| Days between vaccine and CSU* | 10.0 (8.0 – 12.0) | 11 (9.0-13.0) |

<sup>1</sup>n (%); Median (IQR) ; CH : Switzerland, VD : Canton of Vaud
\*In 3 individuals, the days between vaccine and CSU could not be calculated.

**Table 2:** Chronic urticaria (CSU) after COVID-19 Booster (only Moderna and Pfizer) for Switzerland (CH) and Vaud (VD) [from 2022-12-01 to 2022-08-31]

| Brand | VAUD |  |  |  | Switzerland |  |  |  |
| --- | --- | --- | --- | --- | --- | --- | --- | --- |
|  | Booster VD<sup>1</sup> | CSU-VD<sup>1</sup> | Inc. rate per 100K [95%CI]<sup>2</sup> | RR [95%CI]<sup>3</sup> | Booster CH<sup>1</sup> | CSU-CH<sup>1</sup> | Inc. rate per 100K [95%CI]<sup>2</sup> | RR [95%CI]<sup>3</sup> |
| Pfizer | 141,797 (48) | 3 (4) | 2.1 [0.6 - 6.7] |  | 1,307,837 (40) | 25 (4) | 1.9 [1.3 - 2.9] |  |
| Moderna | 157,016 (52) | 69 (96) | 43.9 [34.5 - 56.0] | 20.8 [6.5 - 66.0] | 1,970,971 (60) | 607 (96) | 30.8 [28.4 - 33.4] | 16.1 [10.8 - 24.0] |
| Overall | 298,813 (100) | 72 (100) | 24.1 [19.0 - 30.5] |  | 3,278,808 (100) | 632 (100) | 19.3 [17.8 - 20.9] |  |

<sup>1</sup>N (%)
<sup>2</sup>Incidence rate per 100'000 persons - 95%CI : 95% confidence intervals
<sup>3</sup>Relative risk (reference : Pfizer)
Sources : Federal Office for Public health, Swissmedic, Chief medical officer Vaud, CHUV, Unisanté

## Discussion

Our results suggest an association between the booster dose, the Moderna vaccine, and the new onset of CSU, although this adverse event remains rarely reported. A confounding association with the Omicron variant wave is possible but only 31% reported a confirmed infection in CSU-VD. As a potential contributing mechanism warranting further investigations, the Moderna vaccine was significantly more associated with positive skin and basophil activation tests against either of the two mRNA-based vaccines[5].

Overall, these data should not discourage patients from being vaccinated, as this vaccination campaign has been instrumental in reducing COVID-19 burden and preventing millions of deaths[6]. However, urgent guidelines defining the eligibility/dosing for upcoming mRNA-based boosters are needed for patients with systemic urticaria after a mRNA-based COVID-19 vaccine.

## Data Availability

All data produced in the present study are available upon reasonable request to the authors

## Acknowledgment

Herein we would like to thank the many allergists who helped in the identification of patients with CSU and in particular Dr. O.Estoppey, Dr S.Petitpierre, Dr V. Morel, Dre A. Borgeat, J. Winter Burdet, Dre F. Langener-Viviani, Dr. S Chappuis, Dr. Alain Mantegani, Dr. J.Ducommun, Dr. Y.Coatternec, Dr. R.Opplinger and Dr.L Debétaz.

---

**Table S1.** Chronic urticaria (CSU) after COVID-19 Vaccine - CSU cases after first booster for Switzerland (CH) and Vaud (VD) [from 2022-12-01 to 2022-08-31]

| Characteristic | CSU-VD, N = 72<sup>1</sup> | CSU-CH, N = 636<sup>1</sup> |
| --- | --- | --- |
| Gender |  |  |
| F | 50 (69) | 361 (57) |
| M | 22 (31) | 263 (41) |
| Unknown | 0 | 12 (2) |
| Age (years) | 41 (36 – 49) | 39 (33 – 47) |
| Unknown | 0 | 86 |
| Covid infection | 22 (31) |  |
| Days between vaccine and COVID infection | 48 (18 – 84) |  |
| CSU symptoms worse after COVID infection | 8 (11) |  |
| CSU active at data collection | 62 (86) | 564 (89) |
| Unknown | 0 | 31 (4.8%) |
| Duration of resolved CSU (days) | 82 (55 – 95) |  |
| Brand vaccine that triggered CSU |  |  |
| Moderna | 69 (96) | 607 (95) |
| Pfizer | 3 (4.2) | 25 (4) |
| Unknown | 0 | 4 (1) |
| Dose after which CSU appeared |  |  |
| 3 | 72 (100) | 636 (100) |
| Days between vaccine and CSU* | 10.0 (9.0 – 12.0) |  |

<sup>1</sup>n (%); Median (IQR), \*In 3 individuals, the days between vaccine and CSU could not be calculated.

